# Epidemic meningococcal meningitis outbreak due to serogroup-c in Yobe state northeastern Nigeria: risk factors and effect of reactive ring vaccination with pentavalent conjugate meningococcal vaccine

**DOI:** 10.1101/2025.03.31.25324991

**Authors:** Baba Waru Goni, Hamidu Suleiman Kwairanga, Mohammed Isiaka, Ahmad Ba’aba, Musa Mohammed Baba, Habu Abdul, Muhammad Lawan Gana, Ibrahim Musa Kida, Bukar Bakki, Haruna Yusuph, Mahmoud Bukar Maina, Idris Mohammed

## Abstract

**Background:** There had been a lull in the outbreak of epidemic meningococcal meningitis in Yobe State NE Nigeria for over a decade following the introduction of conjugate monovalent meningococcal vaccine against serogroup-A meningitis. However, an outbreak of serogroup C meningococcal meningitis was recorded for the first time in the state in 2024.

**Objective:** to describe the epidemiology of epidemic meningococcal meningitis outbreak due to serogroup C in Yobe State northeastern Nigeria and assess the effectiveness of reactive focused ring vaccination with conjugate pentavalent meningococcal vaccine (Men5) used for the first time in the state and Nigeria in the control of the outbreak. Methodology: cross sectional descriptive epidemiologic study.

**Results:** 2,948 cases of epidemic meningococcal meningitis and 72 mortalities (CFR 2.4%) were recorded between March and June 2024 in 12 (70.4%) out of 17 local government areas of the state. The highest incidence and mortality was recorded in the age group 1-20 years [2,547 (86.4%)], 54 (CFR 2.1%)] respectively, and lowest in the age group > 40 years [43 (1.5%), 0 (CFR 0.0%)]. Reactive ring vaccination with Men5 was administered to 581, 025 persons aged 1-30 years. There was a significant declined in both the incidence and mortality due to meningococcal meningitis following focused reactive ring vaccination campaign. Few [223, (0.0038%)] mild adverse events following immunization (e.g. mild to moderate pain or swelling or itching at vaccination site, mild headache, fever etc.) were observed among the vaccinees.

**Conclusion:** The epidemic of meningococcal meningitis due to sero-group C had similar clinical and epidemiologic pattern with previous outbreaks due to serogroup A. The used of mass reactive ring vaccination with Men5 focused on affected communities was both effective and safe in controlling the outbreak as well as preventing mortality due to meningitis in Yobe State northeastern Nigeria.

## INTRODUCTION

Epidemic meningococcal meningitis also known as Cerebrospinal Meningitis (CSM) is caused by a gram-negative diplococcal bacterial pathogen called *Neisseria meningitidis*. It is specific to humans and responsible for the outbreak of meningococcal meningitis particularly in the African meningitis belt [1,2]. Twelve different serogroups of *N.meningitidi*s have been established based on the differences in the composition of the surface bacterial polysaccharide capsule. However, serogroups A, B, C, W, Y and X are known to be associated with invasive meningococcal disease (IMD) resulting in both sporadic and epidemic cases of meningococcal meningitis [1,2]. The endemicity of some of these serogroups particularly in the African meningitis belt has been responsible for the cyclical epidemic of meningococcal meningitis in the region.[1,2]

The meningitis belt of Africa was first described by a French Epidemiologist Léon Lapeyssonnie in 1963.[3] It is an area lying between latitudes 4^0^ and 16^0^ North of the equator stretching from Senegal and the Gambia on the West to Ethiopia and Somalia on the East. It is a region characterized by high incidence of cyclical epidemics of meningococcal meningitis and semi-arid climate with scanty average annual rainfall of isohytes (300-1100mm).[3,4,5] Documented evidence of outbreaks of meningitis in the African Meningitis Belt had been reported between 1905 and 1908 and since then cyclical epidemics of meningococcal meningitis occur in the region every 5-10 years. [6] The belt is mainly located in sub-Saharan Africa and is dry and dusty for most of the year. The ‘Harmattan’ season from November to February is characterized by dusty, dry and cold northeasterly wind that originates from the Sahara Desert; is one of the drivers of this epidemic.[4] Epidemics in the African Meningitis Belt were primary caused by *Neisseria meningitidis* sero-group A (NmA).[7,8] The epidemic usually comes in large scale outbreaks in which tens of thousands of people are affected and thousands of deaths recorded every 5-10 years. The original description of the African Meningitis Belt by Lapeyssonnie was restricted to most of the Sahelian parts of West and Central Africa including Sudan, Ethiopia, Djibouti and Somalia. Although countries in the Sahel bear the greatest brunt of the epidemic; subsequent World Health Organization (WHO) surveillance system report have shown that the area has now extended beyond the borders originally described by Lapeyssonnie.[6,7]

Polyvalent polysaccharide vaccines had been used for the control of outbreaks of meningococcal meningitis in Africa for over 3 decades.[8–11] However, there were a lot of drawbacks in the used of polysaccharide vaccine for mass vaccination campaigns during outbreaks of meningococcal meningitis. These include among other things poor induction of immunologic memory particularly in young children[12,13] and logistic problems in maintaining the supply chain for reactive vaccination campaigns when epidemics were underway.[14] One of the major outbreaks of meningococcal meningitis in the African Meningitis Belt was caused by sero-group A in 1996 in northern Nigeria and Niger Republic in which about 250,000 cases were reported and 25,000 deaths recorded.[15,16]

However, since the introduction of conjugate monovalent vaccine against sero-group A (MenAfriVac™) in 2010 in the region, the overall incidence of epidemic meningococcal meningitis had declined steadily along with the risk of its outbreaks. [17] The decrease in the incidence of meningococcal meningitis in the African meningitis belt was largely due to mass vaccination campaigns following the introduction of the MenAfriVac™ vaccine in the region. Sero-group A (NmA) *Neisseria meningitis* cases have disappeared completely in most countries, with sporadic cases reported among unvaccinated individuals in Burkina Faso, Cameroon, Chad, Guinea, Niger, Nigeria, Senegal, Guinea, and Nigeria.[17–20] A rapid herd immunity effect was shown to be statistically significant, leading to a risk reduction for meningitis in the unvaccinated population younger than 1 year and older than 30 years that were not eligible for vaccination. [17–20]

The novel pentavalent (Men5) conjugate meningococcal vaccine is a game changer in the fight against epidemic meningococcal meningitis. It is both safe and effective against the various serogroups of *N. meningitidis* including the evasive X serogroup as demonstrated in several clinical trials thereby giving hope for possible eradication of meningitis.[21] The WHO prequalified this vaccine based on these trials and subsequently recommended its integration into the routine immunization programs in countries of the African meningitis belt. Nigeria has set a record as being the first country in the African meningitis belt and indeed the world to roll out this new vaccine. The support for this new vaccine roll-out by development partners like the Global Alliance for Vaccines Immunization (GAVI), Vaccine Alliance etc underscores the importance of coordinated international efforts in the fight against infectious diseases with epidemic potentials, which aligns with the WHO’s roadmap for the elimination of meningitis by 2030. [21]

The objective of this study was to describe the epidemiology of meningococcal meningitis outbreak due to serogroup-C (NmC) in 2024 in Yobe State, north eastern Nigeria, and to appraise the impact of the newly introduced Men5 vaccine in controlling the outbreak. The new Men5 vaccine was introduced for the first time in Nigeria and Yobe State as well. Yobe State lies within the African meningitis belt; where change in serogroup patterns had been anticipated, including the emergence of serogroup C in recent times. The recent emergence of epidemic meningococcal meningitis due to sero-group C in the state poses a new public health challenge. It is important therefore, to understand the epidemiology of the outbreak and the effectiveness of Men5 in the control of the outbreak as well as improving response and prevention for future epidemics.

The Nigeria Centre for Disease Control (NCDC) gave a case definition for meningitis cases based on clinical and laboratory <u>criteria</u>. A *suspected case* involves sudden fever onset (>38.5°C rectal or >38.0°C axillary) with meningeal signs like neck stiffness, altered consciousness, or bulging fontanelle in toddlers. A *confirmed case* is a suspected or probable case with bacterial pathogens (e.g., *Neisseria meningitidis*, *Streptococcus pneumoniae*, or *Haemophilus influenzae* type b) detected in CSF or blood. A *probable case* has a turbid CSF appearance, high leukocyte count, or bacteria identified by Gram stain. For infants, CSF leukocyte counts >100 cells/mm³ or counts between 10-100 cells/mm³ with high protein or low glucose indicate probable cases. The alert threshold for outbreak detection is three suspected cases per 100,000 people in populations of 30,000–100,000, or two cases in one week for smaller populations, or any unusual case increase compared to non-epidemic years.[22]

## METHODOLOGY

### Study Design

A retrospective cross-sectional descriptive epidemiologic study of the 2024 epidemic meningococcal meningitis outbreak in Yobe State, northeastern Nigeria.

### Data Collection

Data was obtained from the State Epidemiologic Unit in the Public Health Department of the Yobe State Ministry of Health Nigeria, on the 2024 epidemic meningococcal meningitis outbreak.

### Study Area

Yobe State is one of the six states in Northeastern Nigeria, with a projected population of approximately 4.5 million, based on projections from the 2006 Nigerian National Census.[23] It covers a land mass area of 45,502 km² and shares an international boundary with the Diffa and Zinder regions of Niger Republic to the north.

### Ethical Statement

The study was approved by the Health Research and Ethics Committee (HREC) of the Yobe State Ministry of Health, Nigeria (Approval No: MOH/GEN/747) dated 28^th^ October, 2024 and the data was accessed on 6^th^ November, 2024. Confidentiality was strictly maintained for all data used, and individuals whose data were included in the study remain anonymous. Due to the retrospective nature of the study and the use of anonymized patient records, the requirement for informed consent was waived by the ethics committee. No personally identifiable information was collected or used in the study.

## RESULTS

A total of 2,948 cases of meningococcal meningitis were reported during the outbreak. Among these, 1,596 (54.2%) were males, with a case fatality rate of 3.4% (54 deaths) and 1,244 (42.2%) females with case fatality rate of 1.4% (18 deaths). By Month (Figure 1A), the highest number of cases were recorded in March [1,159 (39.3%)], with 34 mortalities (CFR 2.9%), and the lowest recorded in June [41 (1.4%), with no mortality (0.0%). The outbreak affected various age groups (Figure 1B), with the highest number of cases reported in the age group 1-20 years [2,547 (86.4%)], with 54 mortality (CFR-2.1%). In contrast, the lowest number of cases was recorded in the age group >40 years [43 (1.5%)], with 0 (0.0%) mortality. Comparing cases between urban and rural areas (Figure 1C): 1,989 cases (67.5%) were recorded in rural areas and 959 (32.5%) in urban.

**Figure 1A:**
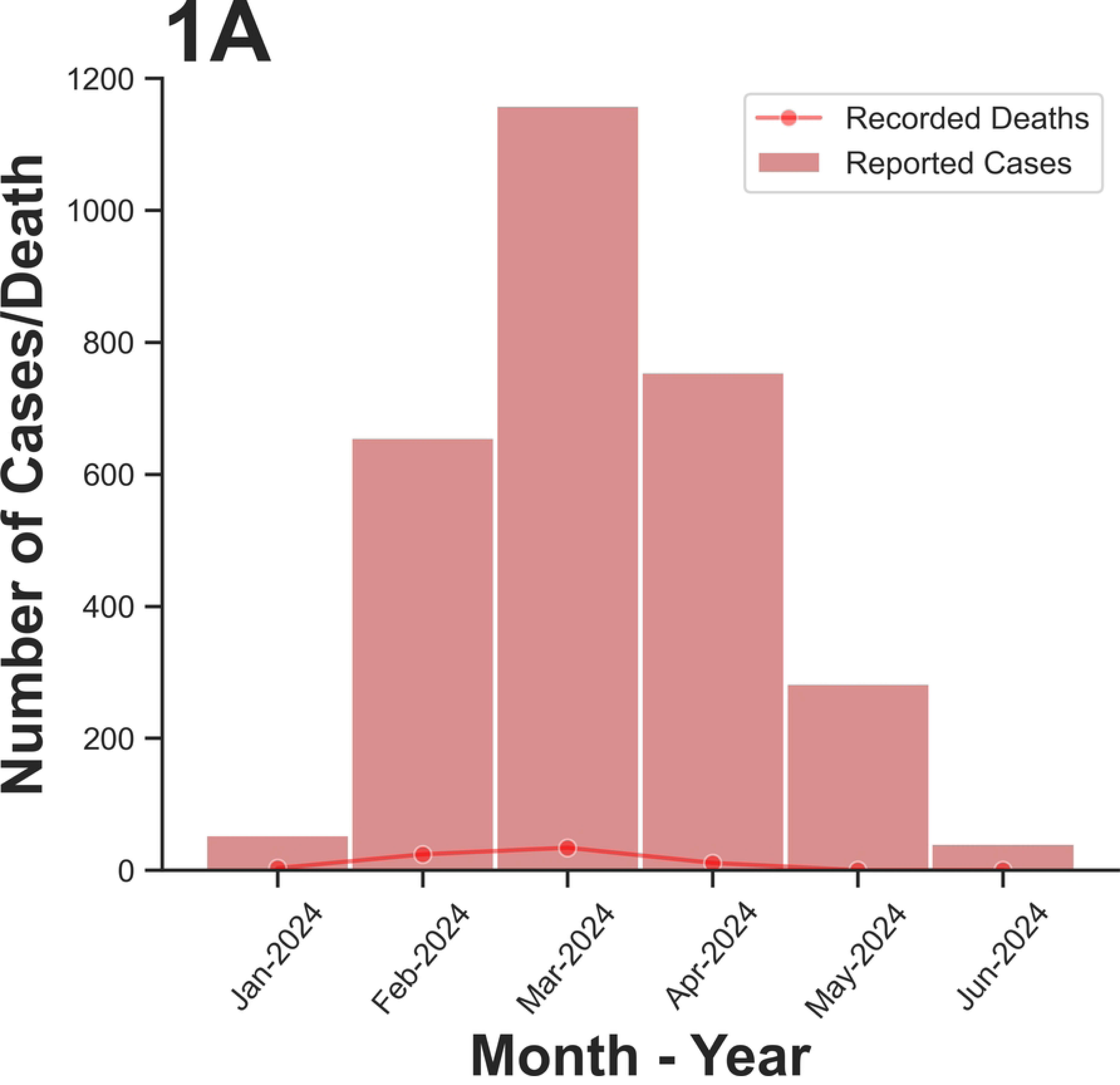
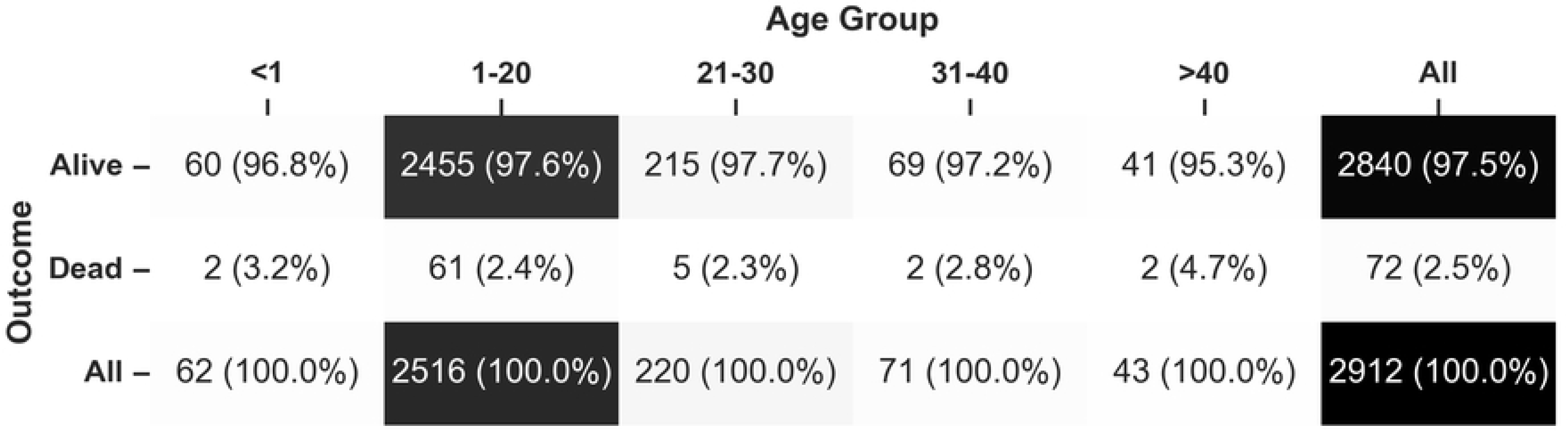
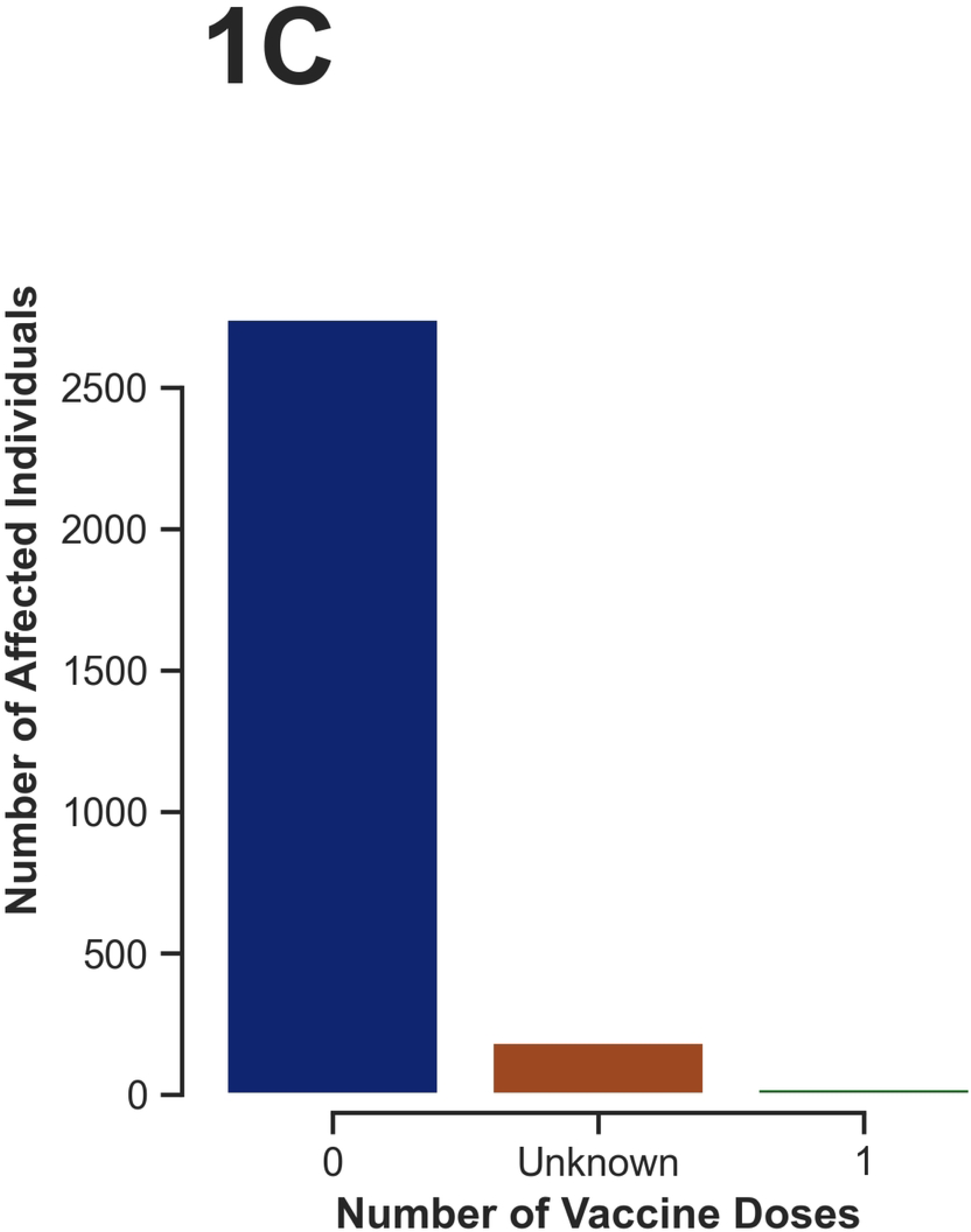
Number of Reported cases/ Recorded death due to meningococal meningitis by month 1B: Heat plot showing the showing the number of reported case/recorded death among different age groups. 1C: Histogram showing the number of reported case/recorded death among Rural/Urban setlers.

Case distribution by local government areas (LGA) was shown in Figure 2A. Meningococcal meningitis outbreak were reported in 12 out of 17 LGAs in the state (70.6%). The highest number of cases were recorded in Potiskum LGA, 780 (26.5%), 17 mortality (CFR-2.1%). Conversely, the lowest was in Bade LGA; 4 (0.1%), 1 mortality (CFR-25%).

**Figure 2A:**
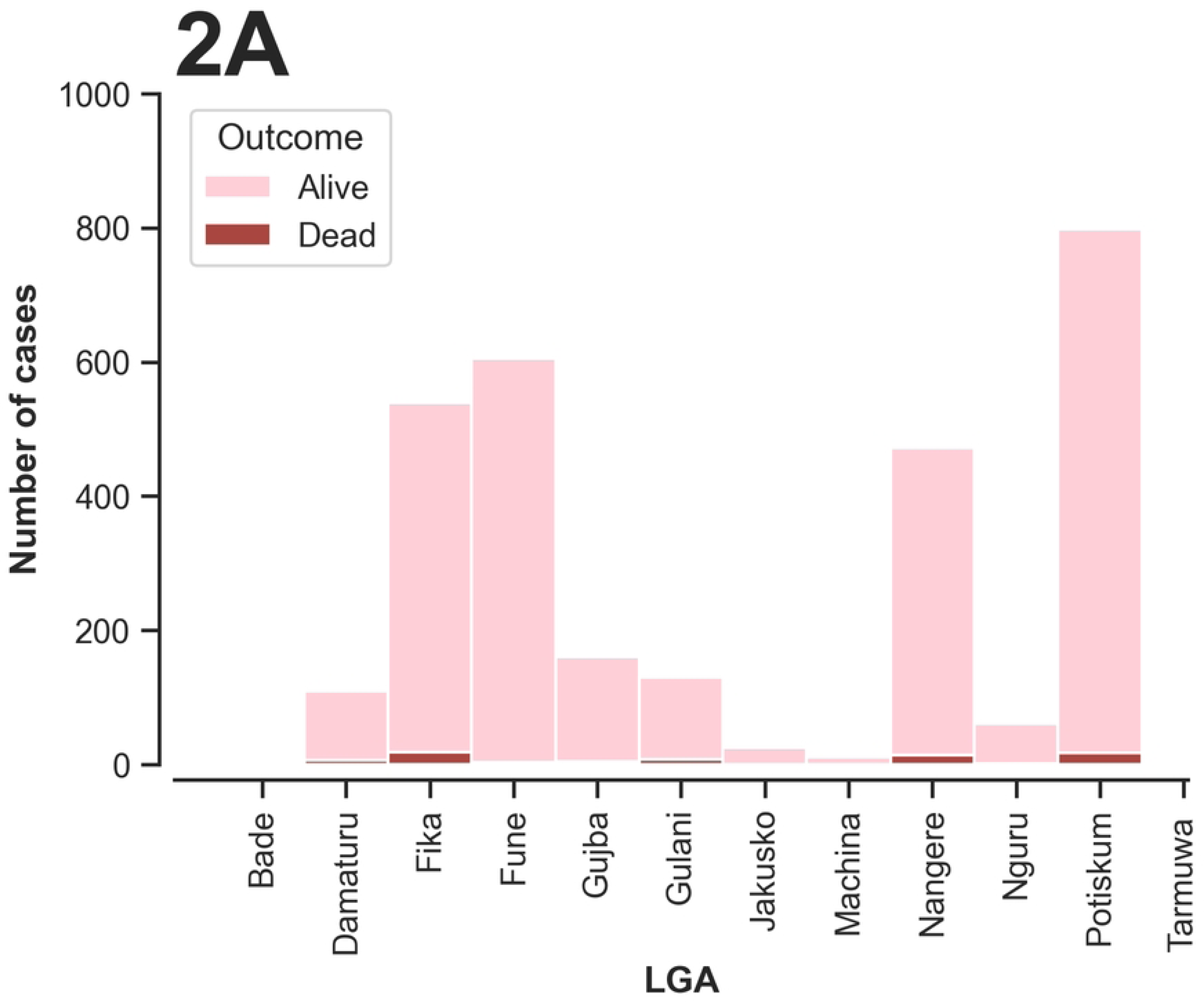
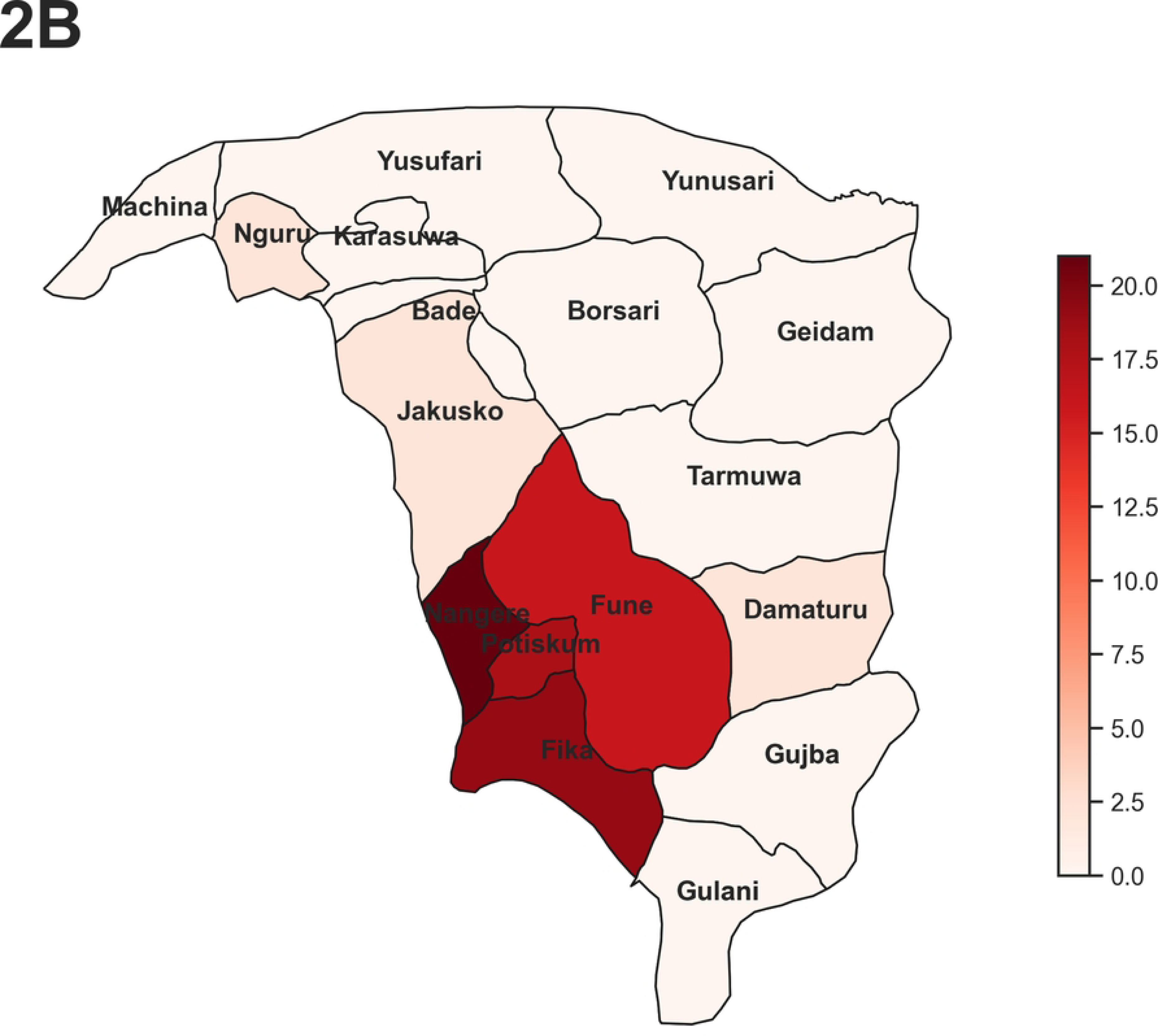
Histogram showing the number of reported case/recorded death due to meningococal meningitis by LGA. **2B:** choropleth map showing the prevalence of meningococal meningitis per 10,000 individuals by LGAs in Yobe state during the 2024 outbreak.

(Figure 2B): shows incidence of epidemic meningococcal meningitis per 10,000 population by local government areas in the state; highest incidence was recorded in Nangere LGA [21.0 per 10,000], followed by Fika LGA [19.0 per 10,000], Potiskum LGA [18.0 per 10,000], Fune LGA [16.0 per 10,000], Gujba LGA [7.0 per 10,000], and Gulani LGA [6.0 per 10,000]. However, the remaining LGAs had incidences of 5.0 per 10,000 or less.

Based on the clinical case definition: (Figure 3B), the most commonest symptom was fever, 412 (99.51%), followed closely by headache, 381 (91.97%), while the least was convulsion or loss of consciousness; 102 (24.63%). In terms of symptom combinations among those diagnosed by clinical case-definition; combination of 4 symptoms viz; fever, headache, neck stiffness and abdominal pain was the commonest [96 (23.17%)], while the least was the combination of fever, headache, neck stiffness, abdominal pain and convulsion / loss of consciousness [1 (0.24%)].

**Figure 3A.**
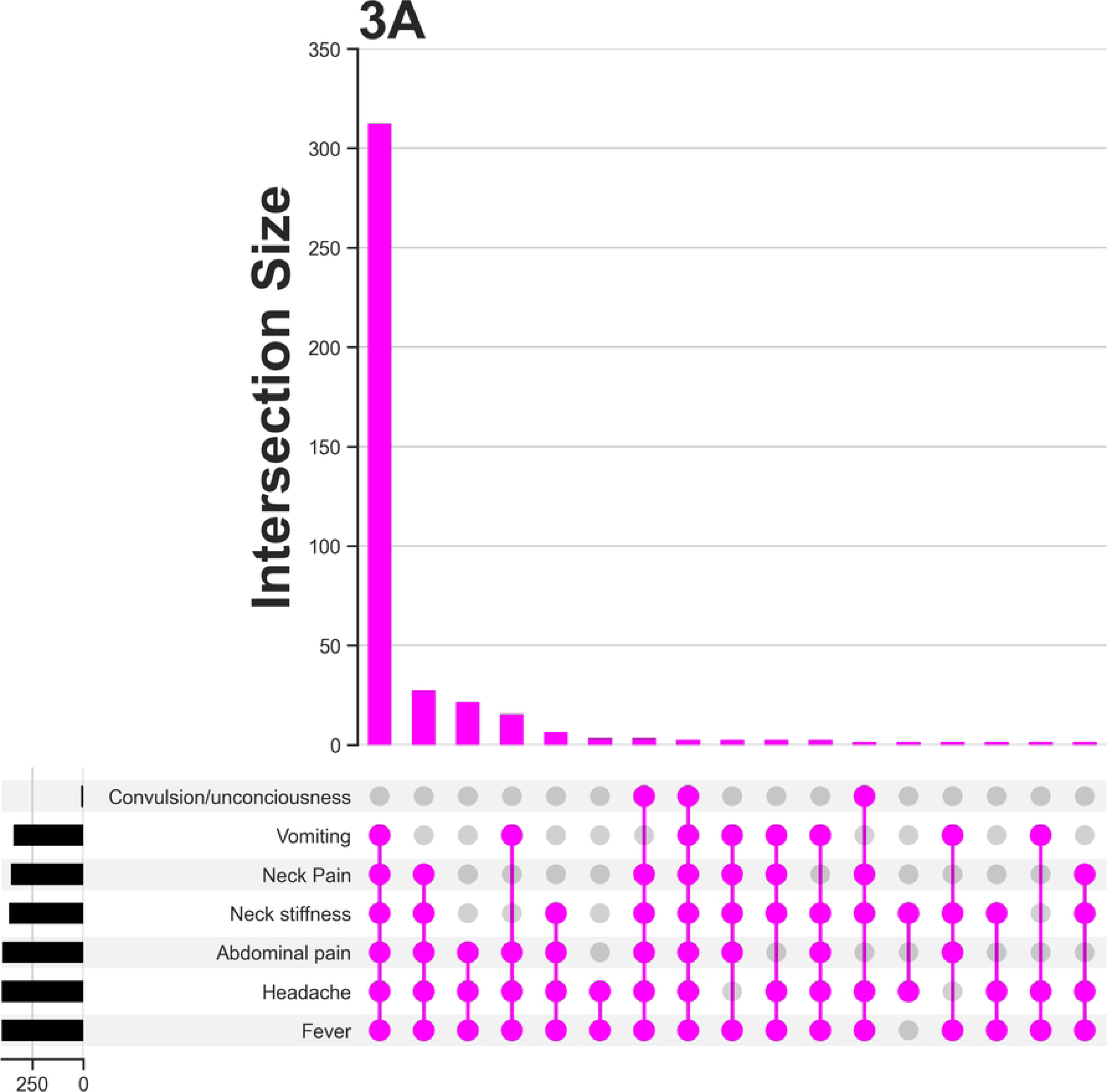
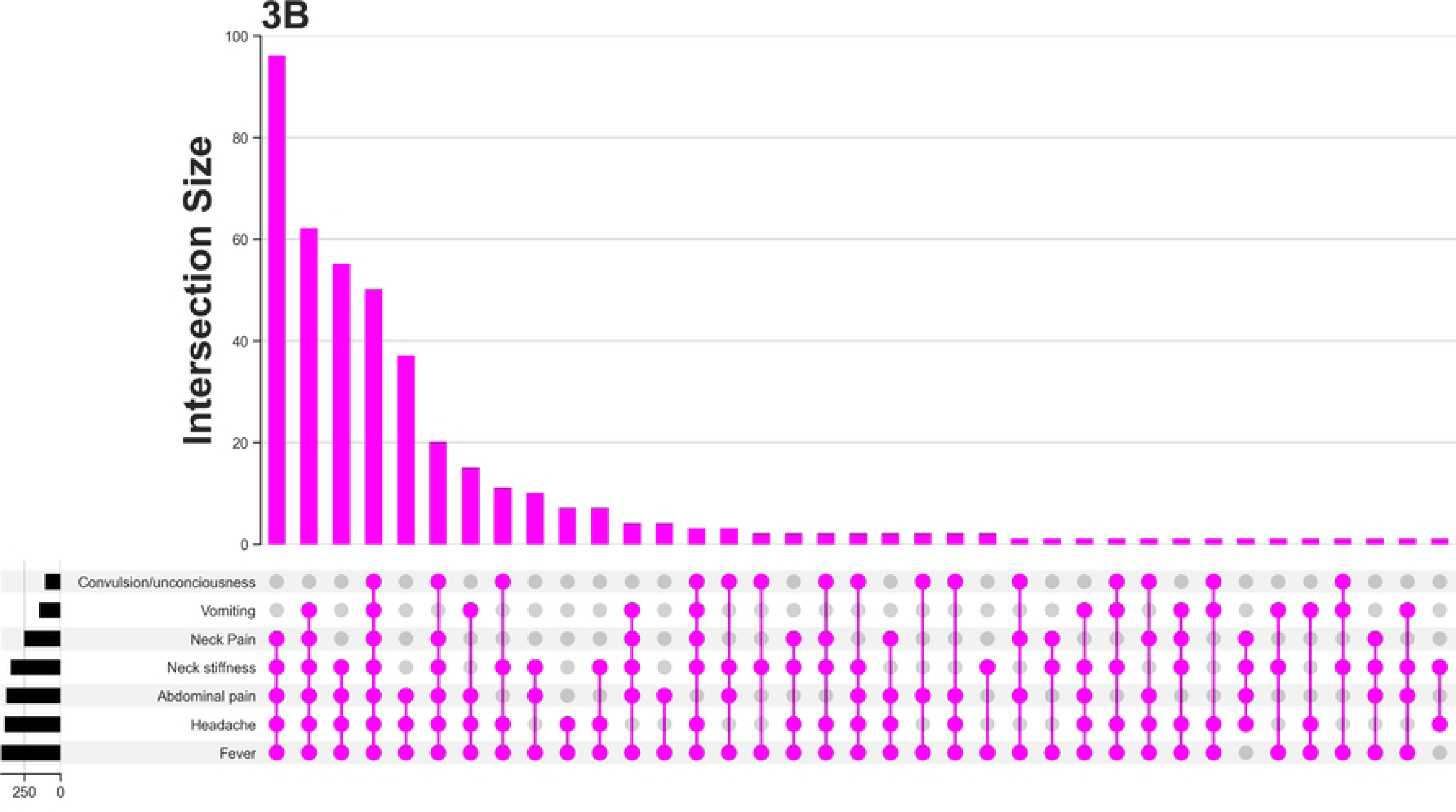
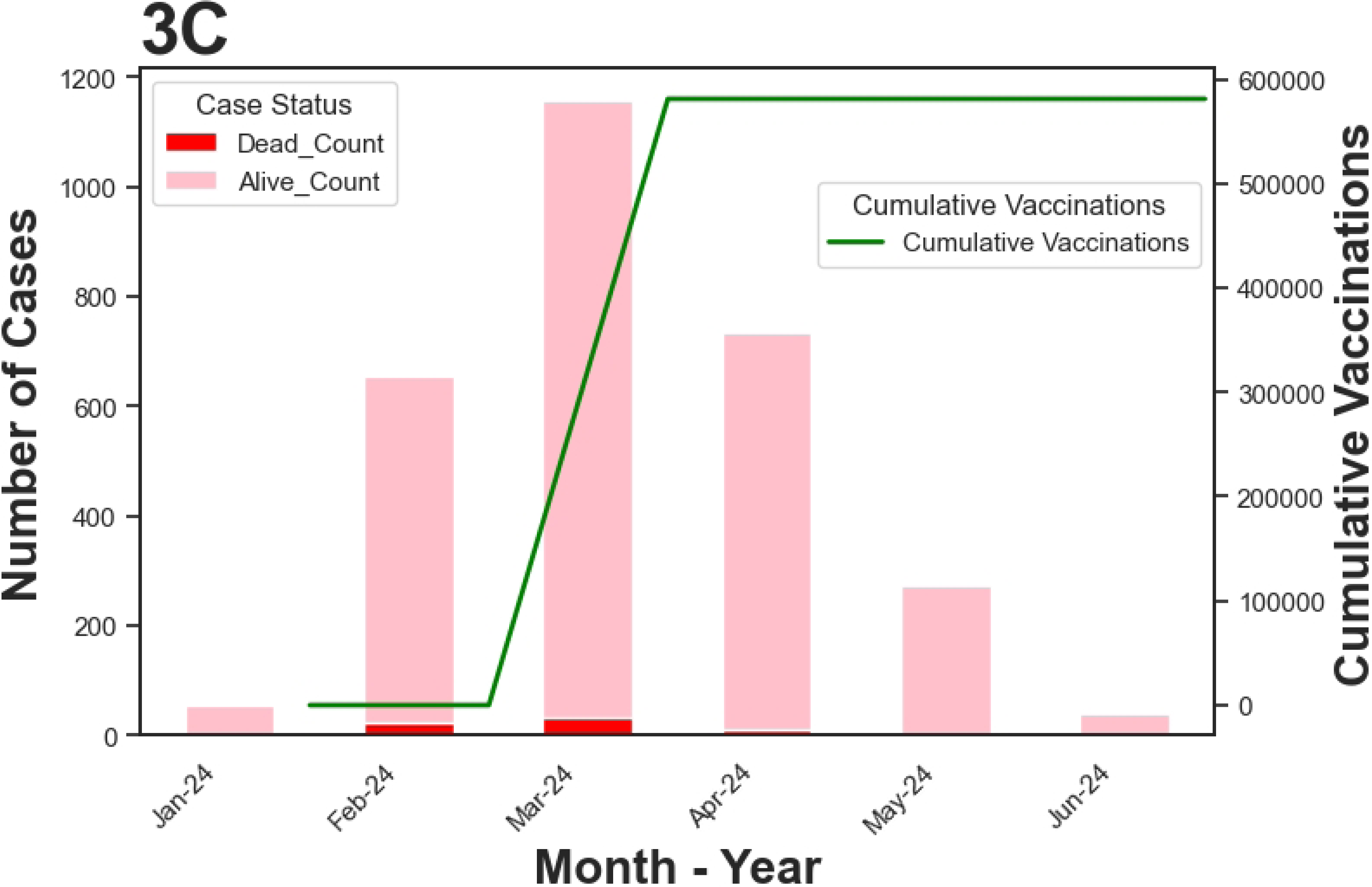
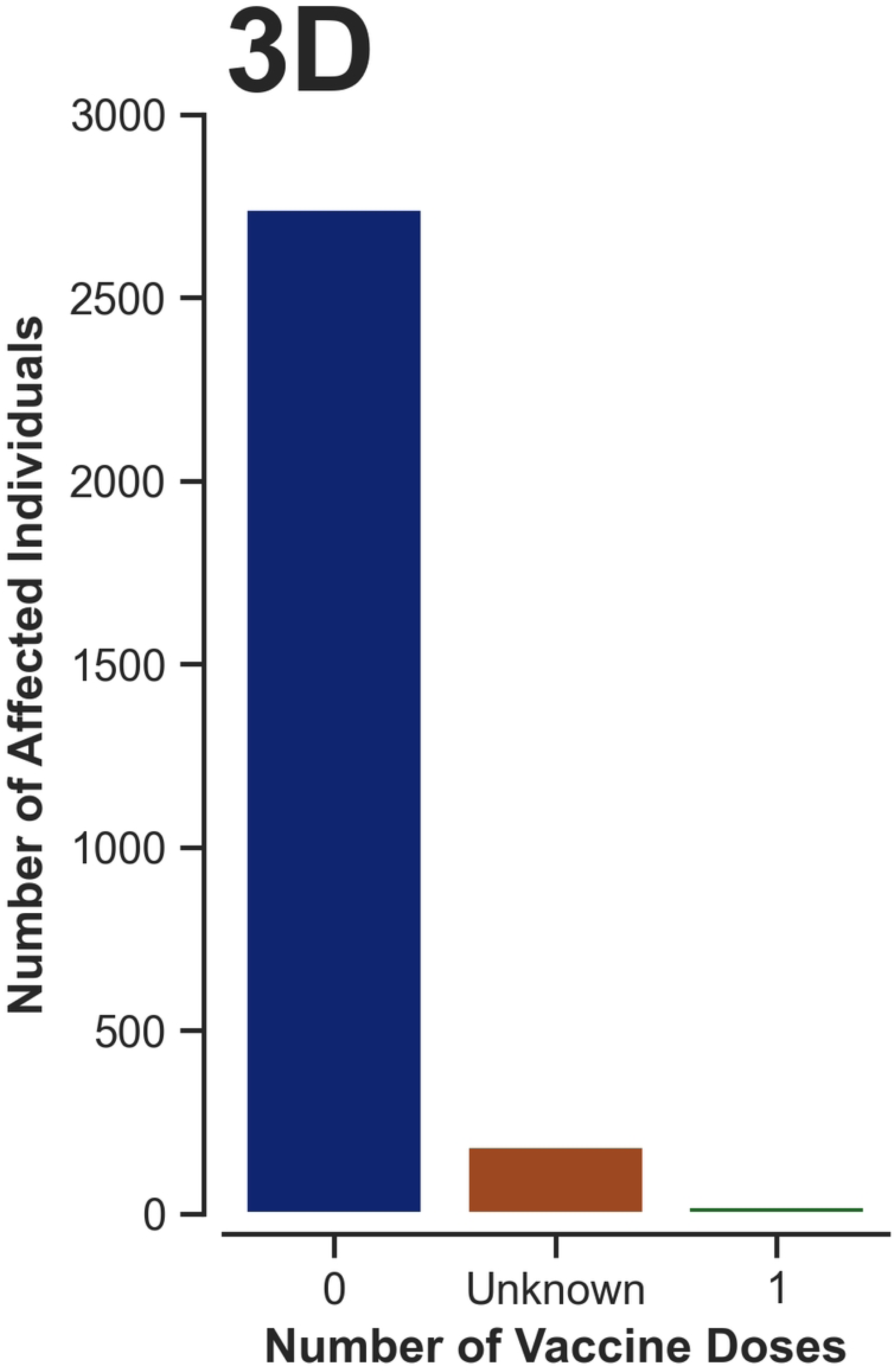
UpSetplot showing combination of symptoms among the lab confiremed cases of meningococal meningitis during the 2024 outbreak in Yobe, Nigeria **3B:** UpSetplot showing combination of symptoms among the total cases of meningococal meningitis during the 2024 outbreak in Yobe, Nigeria **3C:** Combined bar and line plot showing the impact of ring vaccination on number of reported cases/ recorded death during the 2024 outbreak in Yobe, Nigeria **3D:** Histogram showing patients’ vaccination status during the 2024 outbreak in Yobe, Nigeria.

Similarly, among the 86 laboratory confirmed cases (Figure 3A), the most commonest symptom was fever [85, (98.84%)] and the least was convulsion and/or loss of consciousness [ 6 (6.98%)]. In terms of symptoms combination among this group: combination of fever, headache, and abdominal pain was the commonest [21 (24.42%)], while combination of headache and neck stiffness was the least [1 (1.16%)]. A Cohen’s Kappa value of +0.423 indicates a positive correlation between the clinically case defined and laboratory confirmed cases.

A large-scale focused ring vaccination campaign with conjugate pentavalent (Men5) vaccine was commenced in March 2024 in which 581,025 doses were administered to persons 1-30 years old in communities were outbreak occurred (Figure 3C). This was followed by a sudden decline in both the incidence and mortality due to meningitis from a peak of 1,159 in March to 755 in April, representing a 34.87% reduction. This downward trend continued with cases falling further by 62.52% (283 cases) and 85.51% (41 cases) in May 2024 and June 2024 respectively.

The distribution of cases by the number of MenAfricaVac ™ vaccine doses received (against serogroup A meningitis) (Figure 3D) revealed that 99.3 % of cases, including those that died, either had zero-dose or unknown vaccination status (absence of vaccination card for MenAfriVac™) against serogroup A meningitis.

Of the 581,025 persons that received the Men5 vaccine against serogroup-C meningitis (i.e. focused ring vaccination), few [223, (0.038%) had mild adverse events following immunization (AEFI) e.g. mild to moderate pain, swelling or itching at vaccination site, mild headache, fever etc. However, none of the vaccinees had any serious AEFI (e.g. convulsion, loss of consciousness, anaphylactic shock, death etc.).

## DISCUSSION

The 2024 epidemic meningococcal meningitis outbreak in Yobe State northeastern Nigeria revealed gender disparity among cases, this finding was similar to those reported in previous studies suggesting that males may be more exposed to risk factors of IMD than females e.g. local mass gatherings in communities e.g weekly market or worship center gatherings in villages and crowded local public transports in rural areas (e.g. lorries, trucks etc). [27,28] Other risk factors for acquiring nasopharyngeal carriage of meningitis e.g. cigarette smoking and exposure to smoke from wood fires (e.g. bush burning) are common among males.[2] Mortality was also relatively higher in males than females. The outbreak peaked in the month of March 2024 a finding that is in concordance with those reported in similar studies showing the peak incidence of meningitis during the hottest and driest month of the year when conditions such as low relative humidity, high atmospheric temperature and dust storms may increase nasopharyngeal carriage rate and subsequent meningococcal meningitis outbreak susceptibility.[1,2] The lowest number of cases in the month of June may be due to intervention with Men5 vaccine as well as the beginning of the rainy season. This finding is also consistent with findings in similar studies suggesting that the incidence of epidemic meningococcal meningitis declines with the commencement of the rains.[28–30]

Age-group distribution of cases in this study revealed high incidence of cases among younger persons which corroborates with findings reported in other studies indicating that younger individuals are more susceptible meningococcal meningitis than older persons. A plausible explanation could be low immunity as well as increased exposure to risk factors among younger persons (e.g. household crowding or overcrowding in boarding schools).[31,32] In contrast, there were few cases and mortality among the older age group; this may be due to immunity to *Neisseria meningitidis* acquired over time as a result of prior or repeated exposure in the past. Geographically, rural areas accounted for higher incidence of meningitis than urban areas in the study. This rural-urban variation in incidence and mortality due to meningitis is in tandem with findings in other studies that revealed greater vulnerability of rural communities to meningococcal meningitis outbreak due to limited access to preventive healthcare services such as vaccination, risk communication, nutrition etc. People in rural areas are also more exposed to risk factors like smokes from woodfires (e.g. bush burning), wood fuels for cooking as well as intense effects of the Harmattan weather. A possible reason could be poor disease surveillance and delayed outbreak response in hard -to-reach rural communities. Nomadic rural populations with poor access to healthcare services due to their constant geographic mobility may also introduce meningitis into vulnerable sedentary agricultural communities. [30,33,34]

In addition, greater proportion of cases of meningococcal meningitis in this study had zero-dose or unknown status for vaccination against serogroup A vaccination. Vaccination card retention for MenAfriVac™ was also very poor among the study participants, this could be due to ignorance of parents or caregivers on the importance of retaining the immunization cards of their children. This finding may suggest that unimmunized individuals against NmA may be at risk of NmC meningitis. This finding may be due to cross-protection of NmA vaccine against NmC meningitis or due an epiphenomenon. Further studies may be needed to elucidate this finding.

The positive correlation between clinical and laboratory-confirmed cases further reinforces the the need for the commencement of early treatment based on clinical findings during outbreak to prevent complications includings death. [35–37]

The commencement of focused reactive ring vaccination with Men5 vaccine early in the outbreak may have played a crucial role in controlling the epidemic. The declined in both incidence and mortality due to meningitis following the introduction of the vaccine to affected communities had a significant positive impact in controlling the outbreak. This finding lend credence to the fact that timely and effective intervention while an outbreak is underway may be effective in control of meningitis.[38,39]. The use of reactive ring vaccination with Men5 vaccine during this outbreak may also prevent future outbreak due to sero-group C and other serogroups contained in the vaccine. The above finding coupled with the very few cases of mild AEFI amongst the vaccinees (none serious AEFI) suggest that Men5 may be both safe and effective for the control of oubreak of epidemic meningococcal meningitis even when deployed in large scale for mass reactive vaccination campaigns.

## CONCLUSIONS

The outbreak of serogroup C meningococcal meningitis in Yobe State NE Nigeria in 2024 revealed high incidence of meningitis and mortality among the younger age-group, male gender and in rural areas. The commencement of reactive ring vaccination with Men5 vaccine focused on at risk population in affected communities during the outbreak may have resulted in significant decline in both incidence and mortality due to serogroup-C meningitis. This finding underscores the importance of continuous surveillance, timely and targeted vaccination campaigns with Men5 and other public health strategies to prevent and mitigate future epidemics. Mass vaccination campaigns with Men5 vaccine should be commenced early enough before the dry season for the clearance of nasopharyngeal carriage (which is the main driver for an outbreak). This may allow time for the development of long-lasting immunologic memory among vulnerable population before the dry season. Community-based risk communication strategies during outbreaks may help vaccine uptake and prevent local myths and misconceptions about meningitis. Another important step in curtailing the outbreak of meningitis may include effective surveillance systems to monitor meningitis incidences and mortality in real-time during outbreaks as well as continuous training of healthcare workers to improve the diagnostic accuracy and effective clinical management of meningitis.

## Data Availability

The data is available on a public GitHub repository and the link is shared in the article

https://github.com/shkwairanga1/CSM

## Acknowledgement

We would like to extend our special thanks and appreciation to the Department of Public Health Yobe State Ministry of Health Damaturu Nigeria, the Epidemiology Unit Yobe State Primary Health Care Board and the Africa Field Epidemiology Network (AFENET) Yobe State Field Office Damaturu, for provided the data for this work.

## Data Availability

The raw data, images and other supplimentary materials are available <u>here</u>

## Declaration of conflict of Interest

This study was not sponsored and there is no declaration of interest by any of the authors involved in the study.

## Notes

### Competing Interest Statement

The authors have declared no competing interest.

### Clinical Trial

NA

### Clinical Protocols

https://github.com/shkwairanga1/CSM

### Funding Statement

The study is not funded

### Author Declarations

The study was approved by the Health Research and Ethics Committee (HREC) of the Yobe State Ministry of Health, Nigeria (Approval No: MOH/GEN/747) dated 28th October, 2024 and the data was accessed on 6th November, 2024

## REFERENCE

1. Cooper L V, Robson A, Trotter CL, Aseffa A, Collard J, Daugla DM, et al. Risk factors for acquisition of meningococcal carriage in the African meningitis belt. Tropical Medicine & International Health. 2019;24: 392–400.

2. Trotter CL, Greenwood BM. Meningococcal carriage in the African meningitis belt. Lancet Infectious Diseases. 2007;7: 797–803. doi:10.1016/S1473-3099(07)70288-8

3. Lapeyssonnie L. Cerebrospinal meningitis in Africa. Bull World Health Organ. 1963;28: 1–114.

4. Bauer SE, Im U, Mezuman K, Gao CY. Desert Dust, Industrialization, and Agricultural Fires: Health Impacts of Outdoor Air Pollution in Africa. Journal of Geophysical Research: Atmospheres. 2019;124: 4104–4120. doi:10.1029/2018JD029336

5. Mazamay S, Guégan JF, Diallo N, Bompangue D, Bokabo E, Muyembe JJ, et al. An overview of bacterial meningitis epidemics in Africa from 1928 to 2018 with a focus on epidemics “outside-the-belt.” BMC Infect Dis. 2021;21: 1–13. doi:10.1186/S12879-021-06724-1/FIGURES/5

6. Omeh D, Ojo B, Omeh C. Recurring Epidemics of Meningococcal Meningitis in African Meningitis Belt: A Review of Challenges and Prospects. J Adv Med Med Res. 2017;22: 1–12. doi:10.9734/JAMMR/2017/34151

7. Organization WH. Cerebrospinal meningitis in Africa. Weekly Epidemiological Record= Relevé épidémiologique hebdomadaire. 1996;71: 311–312.

8. Sidikou F, Potts CC, Zaneidou M, Mbaeyi S, Kadadé G, Paye MF, et al. Epidemiology of Bacterial Meningitis in the Nine Years Since Meningococcal Serogroup A Conjugate Vaccine Introduction, Niger, 2010–2018. J Infect Dis. 2019;220: S206–S215. doi:10.1093/INFDIS/JIZ296

9. Laval CAB, Pimenta FC, de Andrade JG, Andrade SS, de Andrade ALSS. Progress towards meningitis prevention in the conjugate vaccines era. Brazilian Journal of Infectious Diseases. 2003;7: 315–324. doi:10.1590/S1413-86702003000500006

10. Organization WH. Investing to defeat meningitis and beyond. World Health Organization; 2024.

11. Joshi VS, Bajaj IB, Survase SA, Singhal RS, Kennedy JF. Meningococcal polysaccharide vaccines: A review. Carbohydr Polym. 2009;75: 553–565. doi:10.1016/J.CARBPOL.2008.09.032

12. MacDonald NE, Halperin SA, Law BJ, Forrest B, Danzig LE, Granoff DM. Induction of Immunologic Memory by Conjugated vs Plain Meningococcal C Polysaccharide Vaccine in Toddlers: A Randomized Controlled Trial. JAMA. 1998;280: 1685–1689. doi:10.1001/JAMA.280.19.1685

13. Bugya Z, Prechl J, Szénási T, Nemes É, Bácsi A, Koncz G. Multiple Levels of Immunological Memory and Their Association with Vaccination. Vaccines 2021, Vol 9, Page 174. 2021;9: 174. doi:10.3390/VACCINES9020174

14. Kumar R, Srivastava V, Baindara P, Ahmad A. Thermostable vaccines: an innovative concept in vaccine development. Expert Rev Vaccines. 2022;21: 811–824. doi:10.1080/14760584.2022.2053678

15. Mohammed I, Nasidi A, Alkali AS, Garbati MA, Ajayi-Obe EK, Audu KA, et al. A severe epidemic of meningococcal meningitis in Nigeria, 1996. Trans R Soc Trop Med Hyg. 2000;94: 265–270. doi:10.1016/S0035-9203(00)90316-X

16. Mohammed I, Iliyasu G, Habib AG. Emergence and control of epidemic meningococcal meningitis in sub-Saharan Africa. Pathog Glob Health. 2017;111: 1–6. doi:10.1080/20477724.2016.1274068

17. Bita Fouda AA. Effects of MenAfriVac® Introduction in the African Meningitis Belt, 2010-2017. 2018.

18. LaForce FM, Djingarey M, Viviani S, Preziosi MP. Successful African introduction of a new Group A meningococcal conjugate vaccine: Future challenges and next steps. Hum Vaccin Immunother. 2018;14: 1098–1102. doi:10.1080/21645515.2017.1378841

19. Mueller JE. Long-term effectiveness of MenAfriVac. Lancet Infect Dis. 2019;19: 228–229. doi:10.1016/S1473-3099(18)30725-4

20. Torre G La, Viviani S. Efficacy and Effectiveness of the Meningococcal Conjugate Group A Vaccine MenAfriVac® in Preventing Recurrent Meningitis Epidemics in Sub-Saharan Africa. Vaccines 2022, Vol 10, Page 617. 2022;10: 617. doi:10.3390/VACCINES10040617

21. Ukoaka BM, Okesanya OJ, Daniel FM, Affia MO, Emeruwa VE. A perspective on the novel pentavalent Men5CV (NmCV-5) meningitis vaccine and Nigeria’s pioneering rollout campaign. Infez Med. 2024;32: 323. doi:10.53854/LIIM-3203-6

22. NCDC. Meningitis Fact Sheet. 2019 [cited 3 Nov 2024]. Available: https://ncdc.gov.ng/diseases/factsheet/49

23. NBS. Demographic Statistics Bulletin. 2023 [cited 26 Sep 2024]. Available: https://www.google.com/url?sa=t&source=web&rct=j&opi=89978449&url=https://www.nigerianstat.gov.ng/pdfuploads/DEMOGRAPHIC_BULLETIN_2022_FINAL.pdf&ved=2ahUKEwiaqrDIyeGIAxX1WEEAHb-YAgEQFnoECBcQAw&usg=AOvVaw3X7pva81AWTjAEEy-VeHU0

24. Kehinde OM, Lawan B, Umar AA. Changing Patterns of Temperature in Yobe State, North-Eastern Nigeria: An Evidence of Climate Change. Asian Journal of Geographical Research. 2021; 52–72. doi:10.9734/AJGR/2021/V4I130126

25. Omole O, Welye H, Abimbola S. Boko Haram insurgency: Implications for public health. The Lancet. 2015;385: 941. doi:10.1016/S0140-6736(15)60207-0

26. Suleiman MS, Ajadi SK. Assessment of impact of Boko Haram insurgency on health of internally displaced persons in north east, Nigeria. Igwebuike: An African Journal of Arts and Humanities. 2022;8: 1–23. doi:10.13140/RG.2.2.18277.47848

27. Dias SP, Brouwer MC, Bijlsma MW, van der Ende A, van de Beek D. Sex-based differences in adults with community-acquired bacterial meningitis: a prospective cohort study. Clinical Microbiology and Infection. 2017;23: 121.e9–121.e15. doi:10.1016/J.CMI.2016.10.026

28. Kabanunye MM, Adjei BN, Gyaase D, Nakua EK, Afriyie SO, Enuameh Y, et al. Risk factors associated with meningitis outbreak in the Upper West Region of Ghana: A matched case-control study. PLoS One. 2024;19: e0305416. doi:10.1371/JOURNAL.PONE.0305416

29. Obiakor MO. Weather Variables and Climatic Influence on the Epidemiology of Cerebrospinal or Meningococcal Meningitis. Asian J Med Pharm Res. 2013;3. Available: www.science-line.com

30. Omoleke SA, Alabi O, Shuaib F, Braka F, Tegegne SG, Umeh GC, et al. Environmental, economic and socio-cultural risk factors of recurrent seasonal epidemics of cerebrospinal meningitis in Kebbi state, northwestern Nigeria: A qualitative approach. BMC Public Health. 2018;18: 127–136. doi:10.1186/S12889-018-6196-9/TABLES/2

31. Diallo K, Trotter C, Timbine Y, Tamboura B, Sow SO, Issaka B, et al. Pharyngeal carriage of Neisseria species in the African meningitis belt. Journal of Infection. 2016;72: 667–677.

32. Mueller JE, Yaro S, Ouédraogo MS, Levina N, Njanpop-Lafourcade B-M, Tall H, et al. Pneumococci in the African meningitis belt: meningitis incidence and carriage prevalence in children and adults. PLoS One. 2012;7: e52464.

33. Tanko Umaru E, Nazri Muhamad Ludin A, Rafee Majid M, Sabri S, Moses CP, Enegbuma W, et al. Risk Factors Responsible for the Spread of Meningococcal Meningitis: A Review. International Journal of Education and Research. 2013;1. Available: www.ijern.com

34. Aliyu BMPhD, Abubakar AD. Social Factors and Management of Cerebrospinal Meningitis in Magama Local Government Area of Niger State. JALINGO JOURNAL OF SOCIAL AND MANAGEMENT SCIENCES. 2021;3: 136145–136145. Available: http://oer.tsuniversity.edu.ng/index.php/jjsms/article/view/88

35. Strelow VL, de Miranda ÉJFP, Kolbe KR, Framil JVS, de Oliveira AP, Vidal JE. Meningococcal meningitis: clinical and laboratorial characteristics, fatality rate and variables associated with in-hospital mortality. Archives of Neuro-Psychiatry. 2016;74: 875–880. doi:10.1590/0004-282X20160143

36. Nissa S, Iftiyastuti B, Dwi Rizkita L, Ihsana N, Selohandono A. The Relationship between the Classic Triad of Meningitis and Types of Meningitis. Journal La Medihealtico. 2024;5: 848–856. doi:10.37899/JOURNALLAMEDIHEALTICO.V5I4.1542

37. Akaishi T, Kobayashi J, Abe M, Ishizawa K, Nakashima I, Aoki M, et al. Sensitivity and specificity of meningeal signs in patients with meningitis. J Gen Fam Med. 2019;20: 193–198. doi:10.1002/JGF2.268

38. WHO. In world first, Nigeria introduces new 5-in-1 vaccine against meningitis. 2024 [cited 15 Oct 2024]. Available: https://www.who.int/news/item/12-04-2024-in-world-first--nigeria-introduces-new-5-in-1-vaccine-against-meningitis

39. WHO. WHO Combats Cerebrospinal Meningitis Outbreak in Yobe State, Nigeria | WHO | Regional Office for Africa. 2024 [cited 15 Oct 2024]. Available: https://www.afro.who.int/countries/nigeria/news/who-combats-cerebrospinal-meningitis-outbreak-yobe-state-nigeria

